# Knowledge, Attitude, and Practice of Hand Hygiene among On-Campus Undergraduate Students at the University of Calabar, Nigeria

**DOI:** 10.1101/2025.11.23.25340821

**Authors:** Harrison Oghenevwegba Odibo, Iko-Obong Donatus Edike, Fortune Ita Sunday

**Affiliations:** University of Calabar, Calabar, Nigeria; University Of Uyo, Uyo, Nigeria

**Keywords:** Hand Hygiene, Knowledge, Attitude, Practice, University Students, Nigeria, Public Health

## Abstract

**Background:** Hand hygiene is a cost-effective and critical measure for preventing the transmission of infectious diseases. However, compliance remains a significant challenge, particularly in high-density settings like university hostels. While studies have focused on healthcare workers, data on the hand hygiene (HH) practices of university students in Nigeria are limited.

**Methods:** A descriptive cross-sectional study was conducted among 218 on-campus undergraduate students at the University of Calabar, selected via a multi-stage sampling technique. Data were collected using a pre-tested, semi-structured, self-administered questionnaire. Knowledge, attitude, and practice (KAP) scores were calculated and categorized. Data analysis employed descriptive statistics and chi-square tests using SPSS version 20.0, with a significance level of p < 0.05.

**Results:** The mean age of respondents was 21.72 ± 2.97 years. Although nearly all respondents (99.1%) were aware of hand hygiene, only 22.5% demonstrated ‘good’ knowledge. In contrast, the majority exhibited a positive attitude (93.1%) and adequate self-reported practice (94.9%). The most common reason for non-compliance was forgetfulness (74.3%). A significant association was found between the hall of residence and both knowledge (p=0.021) and attitude (p<0.001).

**Conclusion:** This study reveals a significant gap between awareness and comprehensive knowledge of hand hygiene among students. Despite positive attitudes and self-reported practices, the knowledge deficit and cited barriers like forgetfulness and inadequate facilities highlight the need for targeted interventions. We recommend infrastructural improvements, sustained health education, and the promotion of hand hygiene role models to bridge the knowledge-practice gap.

## 1. Introduction

Hand hygiene is widely recognized as a cornerstone of infectious disease control and an important, cost-effective public health measure [1, 2]. Despite its proven role in reducing the burden of communicable diseases such as diarrhea and acute respiratory infections, compliance with recommended hand hygiene practices remains persistently low across various populations [3].

The university environment, particularly on-campus hostels, presents a unique setting for disease transmission due to high population density, shared living facilities, and extensive social interaction [4]. While substantial research and intervention efforts have been directed towards hand hygiene among healthcare workers [5, 6], there is a relative paucity of studies focusing on university student populations in low-resource settings. In Nigeria, existing KAP studies have predominantly been conducted in the western region and primarily target health professionals, creating a geographical and demographic knowledge gap [7].

Global studies on student hand hygiene have explored habits, environmental barriers, and educational programs. However, the sustainability of these interventions and the impact of specific behavioral cues, such as teacher modeling, are not well-evaluated [8]. Therefore, this study aimed to assess the knowledge, attitude, and practice of hand hygiene among on-campus undergraduate students of the University of Calabar, Nigeria. The findings are intended to identify existing gaps and inform the development of targeted, context-specific health promotion strategies within the university community.

## 2. Methods

### 2.1. Study Design and Setting

A descriptive cross-sectional study was conducted in May 2023 at the University of Calabar, a federal university in Cross River State, South-South Nigeria. The study setting included the university’s on-campus hostels (Halls 2, 4, 5, 8, and 9).

### 2.2. Study Population and Sampling

The study population comprised male and female on-campus undergraduate students. Students who had resided in the hostels for less than one semester, visiting students (“squatters”), and those unwilling to participate were excluded.

The sample size was calculated as 218 using the Cochran formula for a single proportion, assuming a 95% confidence level (Z=1.96), 5% margin of error (d=0.05), and a proportion (p) of 0.83 for good KAP from a previous study [9]. A multi-stage sampling technique was employed:

1. **Stage 1:** Simple random sampling (balloting) to select 5 out of the 8 hostels.
2. **Stage 2:** Simple random sampling to select floors within each chosen hostel.
3. **Stage 3:** Simple random sampling to select rooms on the chosen floors.
4. **Stage 4:** Simple random sampling to select a maximum of 4 students from each chosen room.

### 2.3. Data Collection Tool and Technique

A pre-tested, semi-structured, self-administered questionnaire was used for data collection. The questionnaire was adapted from prior studies and WHO hand hygiene tools. It was pre-tested on 10% of the sample size (22 students) at the University of Cross River State to ensure validity and clarity; these respondents were excluded from the main study. The tool comprised five sections:

- Socio-demographic characteristics.
- Knowledge of hand hygiene (15 items).
- Attitude towards hand hygiene (8 items on a 5-point Likert scale).
- Practice of hand hygiene (20 items).
- Perceived barriers to hand hygiene.

### 2.4. Measurement and Scoring

- **Knowledge:** A total score of 15 was used. Scores of 0–5, 6–11, and 12–15 were categorized as ‘poor’, ‘fair’, and ‘good’ knowledge, respectively.
- **Attitude:** A total score of 28 was used. Scores of 0–13 and 14–28 were categorized as ‘negative’ and ‘positive’ attitude, respectively.
- **Practice:** A total score of 40 was used. Scores of 0–19 and 20–40 were categorized as ‘inadequate’ and ‘adequate’ practice, respectively.

### 2.5. Data Analysis

Data were analyzed using IBM SPSS Statistics for Windows, Version 20.0. Descriptive statistics (frequencies, percentages, means, and standard deviations) were used to summarize variables. Bivariate analysis using the Chi-square test was employed to test for associations between socio-demographic variables and KAP scores. A p-value of less than 0.05 was considered statistically significant.

### 2.6. Ethical Consideration

Ethical approval for this study was obtained from the University of Calabar Research Ethics Committee. Informed written consent was obtained from all participants after explaining the study’s purpose and procedures. Confidentiality was maintained, and participation was entirely voluntary.

## 3. Results

### 3.1. Socio-Demographic Characteristics

A total of 218 students participated, with a response rate of 100%. The mean age was 21.72 ± 2.97 years, and the majority (59.6%) were male. Most respondents were in their first year of study (31.2%) and hailed from the Faculty of Science (87.6%). (Table 1).

**Table 1:** Socio-demographic Characteristics of Respondents (N=218)

| Variable | Category | Frequency | Percentage (%) |
| --- | --- | --- | --- |
| Age Group | $\leq 20$ | 92 | 42.2 |
|  | 21-25 | 101 | 46.3 |
|  | 26-30 | 23 | 10.6 |
|  | 31-35 | 2 | 0.9 |
| Gender | Male | 130 | 59.6 |
|  | Female | 88 | 40.4 |
| Level of Study | 100 | 68 | 31.2 |
|  | 200 | 42 | 19.3 |
|  | 300 | 60 | 27.5 |
|  | 400 | 27 | 12.4 |
|  | 500 | 16 | 7.3 |
|  | 600 | 5 | 2.3 |
| Faculty | Science | 191 | 87.6 |
|  | Arts | 27 | 12.4 |

### 3.2. Knowledge of Hand Hygiene

Although 99.1% (216/218) of respondents were aware of hand hygiene, only 22.5% (49/218) had ‘good’ knowledge. The majority (77.5%) had ‘fair’ knowledge, and no respondent had ‘poor’ knowledge. Most students (86.7%) correctly identified hand hygiene as washing hands with water and soap, and 99.5% acknowledged its importance in disease prevention. However, only 72.9% could demonstrate the correct steps of hand hygiene (Table 2).

**Table 2:** Knowledge of Hand Hygiene among Respondents (N=218)

| Knowledge Item | Response | Frequency | Percentage (%) |
| --- | --- | --- | --- |
| Aware of Hand Hygiene | Yes | 216 | 99.1 |
|  | No | 2 | 0.9 |
| Defines HH as washing with water & soap | Yes | 189 | 86.7 |
|  | No | 29 | 13.3 |
| Aware of the steps of HH | Yes | 169 | 77.5 |
|  | No | 49 | 22.5 |
| Able to demonstrate steps | Yes | 159 | 72.9 |
|  | No | 59 | 27.1 |
| Overall Knowledge Score | Good (12-15) | 49 | 22.5 |
|  | Fair (6-11) | 169 | 77.5 |
|  | Poor (0-5) | 0 | 0.0 |

### 3.3. Attitude towards Hand Hygiene

The vast majority of respondents (98.2%) agreed that handwashing is part of personal hygiene. Overall, 93.1% (203/218) had a positive attitude towards hand hygiene. A significant proportion (65.1%) strongly agreed that training on proper techniques should be organized. The primary barrier to practice was forgetfulness (74.3%), followed by inadequate facilities (22.9%).

### 3.4. Practice of Hand Hygiene

Self-reported practice was high, with 94.9% (207/218) of students categorized as having ‘adequate’ practice. The most consistently practiced behaviors were washing hands after meals (84.9%), before meals (80.3%), and after using the toilet (78.0%). The least practiced behavior was washing hands after every lecture (27.1%) (Table 3).

**Table 3:**
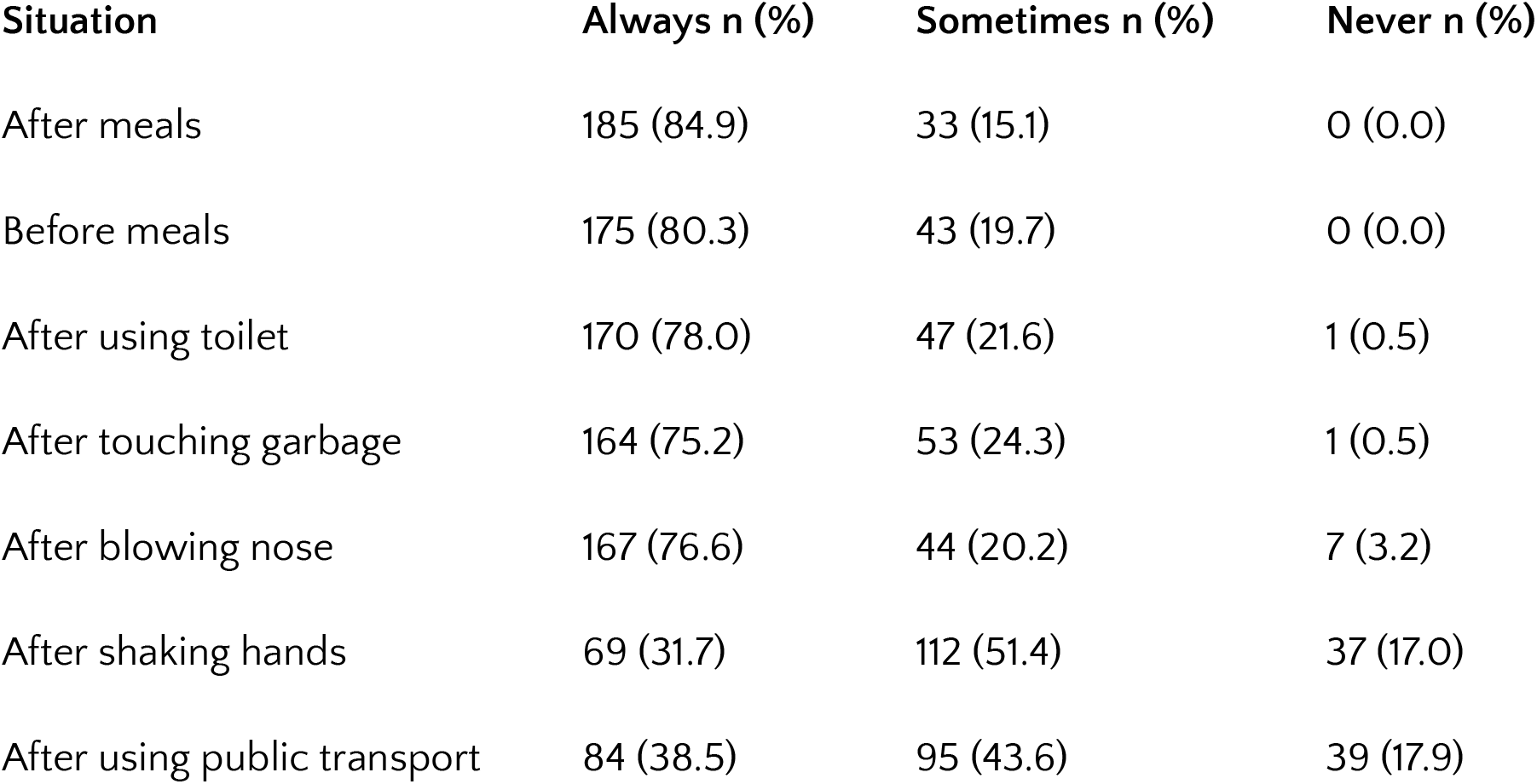
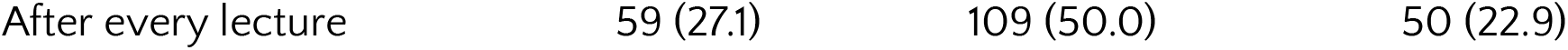
Self-Reported Hand Hygiene Practices among Respondents (N=218)

### 3.5. Association between Socio-demographic Variables and KAP

Bivariate analysis revealed a statistically significant association between the hall of residence and knowledge level (χ^2^ = 7.72, p = 0.021), with students in Halls 2 and 8 more likely to have good knowledge. Hall of residence was also significantly associated with attitude (χ^2^ = 33.60, p < 0.001). No significant associations were found between any socio-demographic variables and hand hygiene practice.

## 4. Discussion

This study provides insight into the hand hygiene KAP of on-campus students at a Nigerian university. The near-universal awareness (99.1%) of hand hygiene is commendable and aligns with studies stressing the penetration of public health messages [10]. However, the translation of this awareness into comprehensive knowledge was deficient, with only 22.5% possessing ‘good’ knowledge. This disparity highlights that mere awareness does not equate to a deep understanding of techniques, indications, and principles, a finding consistent with research by Onyedibe et al. in Nigeria, where good knowledge was also low among non-health professionals [11].

The overwhelmingly positive attitude (93.1%) is a strong asset for public health intervention. Students recognized the value of hand hygiene and supported educational initiatives. However, the predominant barrier of “forgetfulness” suggests that hand hygiene is not yet an ingrained, habitual behavior. This points to a classic intention-behavior gap, where positive attitudes fail to materialize into consistent action without environmental cues and reinforcement [12].

The self-reported adequate practice (94.9%) is higher than in some previous studies [13]. Practices were highest around food-related activities and after using the toilet, which are traditionally emphasized behaviors. Conversely, practices were notably low after lectures, shaking hands, and using public transportation—situations involving high-contact surfaces and interpersonal interaction that are significant for respiratory and contact-borne disease transmission. This indicates a specific area for targeted education.

The significant association between the hall of residence and both knowledge and attitude suggests a potential “peer effect” or subcultural influence within living environments, warranting further qualitative exploration. The lack of association with practice, however, implies that the actual performance of hand hygiene may be influenced more by immediate environmental constraints (e.g., water availability at the moment) than by broad demographic factors.

### 4.1. Limitations of the Study

This study has limitations. The cross-sectional design captures a snapshot in time and cannot establish causality. The reliance on self-reported practices is subject to social desirability bias, likely leading to an overestimation of actual practice. The study was also confined to one university, which may limit the generalizability of the findings.

## 5. Conclusion

In conclusion, this study reveals a critical gap between high awareness and sufficient knowledge of hand hygiene among university students. While attitudes are positive and self-reported practices are encouraging, the deficits in knowledge and the reported barriers indicate that students are not fully equipped or enabled to practice optimal hand hygiene consistently. The university environment presents a strategic opportunity to shape lifelong healthy behaviors.

## 6. Recommendations

Based on the findings, the following recommendations are proposed:

1. The University management should prioritize investment in Water, Sanitation, and Hygiene (WASH) infrastructure, ensuring reliable water supply, functional sinks, and provision of soap or alcohol-based hand rubs at key points in hostels and lecture halls.
2. The University Health Service should implement sustained, multi-channel hand hygiene campaigns. These should move beyond awareness to demonstrate correct techniques, emphasize frequently missed opportunities (e.g., after lectures), and use visual aids like posters near washing stations.
3. Academic and administrative staff should be encouraged to act as hand hygiene role models, reinforcing good practice through their own behavior.
4. Future research should employ direct observation to validate self-reported practices and explore the determinants of hand hygiene behavior through mixed-methods approaches.

## Data Availability

All data produced in the present work are contained in the manuscript

## Acknowledgments

The authors would like to thank the study participants for their time and cooperation. We also acknowledge the support from the Department of Community Medicine, University of Calabar.

